# Monitoring the COVID-19 immunisation programme through a National Immunisation Management System – England’s experience

**DOI:** 10.1101/2021.09.14.21263578

**Authors:** Elise Tessier, Julia Stowe, Camille Tsang, Yuma Rai, Eleanor Clarke, Anissa Lakhani, Ashley Makwana, Heather Heard, Tim Rickeard, Freja Kirsebom, Catherine Quinot, Shreya Lakhani, Linda Power, Michael Edelstein, Andy Evans, Mary Ramsay, Jamie Lopez-Bernal, Joanne White, Charlotte Gower, Nick Andrews, Colin Campbell

## Abstract

In England, the National Immunisation Management System (NIMS) has been used to deliver COVID-19 vaccinations across England, monitor vaccine coverage, and assess vaccine effectiveness and safety.

The NIMS was developed by a joint collaboration between a range of health and digital government agencies. Vaccinations delivered at large vaccination sites, pharmacies, hospitals and in primary care are entered on a point of care application which is verified using the unique NHS number in a centralised system containing information for everyone resident and registered with a GP in England. Vaccination details and additional data from hospital and GP records (such as priority groups) are sent to NHS Digital for data linkage. The NIMS constantly receives updated details from NHS Digital for all individuals and these data are provided to Public Health England (PHE) in a secure environment.

PHE primarily use the NIMS for vaccine coverage, vaccine effectiveness and safety. Daily access to individual-level vaccine data has allowed PHE to rapidly and accurately estimate vaccine coverage and provide some of the world’s first vaccine effectiveness estimates.

Other countries evaluating the roll-out and effect of COVID-19 vaccine programmes should consider a vaccine register or immunisation information system similar to the NIMS.

## Introduction

Monitoring the roll out and delivery of vaccine programmes is essential to ensure their success and maximise population protection against vaccine-preventable diseases^1 2^. The United Kingdom (UK) was among the first countries in the world to roll-out a COVID-19 immunisation programme and has learnt many lessons throughout its roll-out that are valuable to many other countries around the globe.

In England, vaccinations are recorded in the patients General Practice (GP) record and for those delivered to children under 19 years, in Child Health Information Systems (CHIS), a series of sub-national vaccine registers^3 4^. Successful monitoring relies on correct coding at the GP level and on vaccines delivered outside of the GP (for example, in pharmacies, schools, maternity units) being appropriately coded and transferred to the GP record. Research shows that national vaccine registers are key when it comes to monitoring vaccination programmes^1^. The National Immunisations Management System (NIMS) was commissioned by the NHS to improve data flows for the national influenza programme, however, with the urgency and the scale of the COVID-19 vaccination programme (the largest immunisation programme in British history) it was clear that in order to reach the entire population, vaccines would need to be delivered in multiple settings such as hospitals, general practices (GP), pharmacies, mobile vaccination units (e.g. for delivery in care homes), and mass immunisation sites. Furthermore, it was essential to ensure that information on vaccinated individuals was recorded in as near to real time as possible and vaccine coverage data at the national or subnational level, according to groups of interest, was accessible in a timely, complete and accurate manner. Consequently, the NIMS was built by a range of health and digital government agencies^5 6^. Since autumn 2020 the NIMS has been used to collect vaccination information at the individual level in a centralised system for the management of both seasonal influenza and COVID-19 vaccinations across England.

This paper describes the development of the NIMS and how it is used to estimate vaccine coverage and monitor vaccine effectiveness and safety in England. This paper also validates the NIMS data using questionnaire data used for enhanced surveillance purposes.

## Methods

### Development and use of the NIMS

The NIMS has multiple functions, including identifying priority groups eligible for COVID-19 vaccines, sending invitations and reminders for vaccination appointments, and monitoring those invited for a vaccination and recording vaccination data^7^. Currently there are 3 main point of care applications (PoC apps) available and standardised for any vaccination setting to collect vaccination details on Individuals who have presented to a vaccination site (e.g. GP, pharmacy, hospital provider, or mass immunisation site) to receive a COVID-19 vaccine. Several data items are mandatory to enter in the PoC app, such as the unique patient identifier NHS number, date of vaccination and batch number. The NHS number are entered on the PoC apps are verified with the NHS Spine (a centralised IT infrastructure allowing NHS services to exchange of information across local and national NHS) for everyone resident and registered with a GP in England to ensure completeness and accuracy^8^.

NHS Digital, the UK government’s Digital Health Agency, validate and link the data from GP and hospital records to identify individuals belonging to different cohort groups. NHS Digital then send this data through to the NIMS and to the individual’s GP record. Public Health England (PHE) receives these data daily from the NIMS in a secure Azure environment which is then stored in a SQL database (Figure 1).

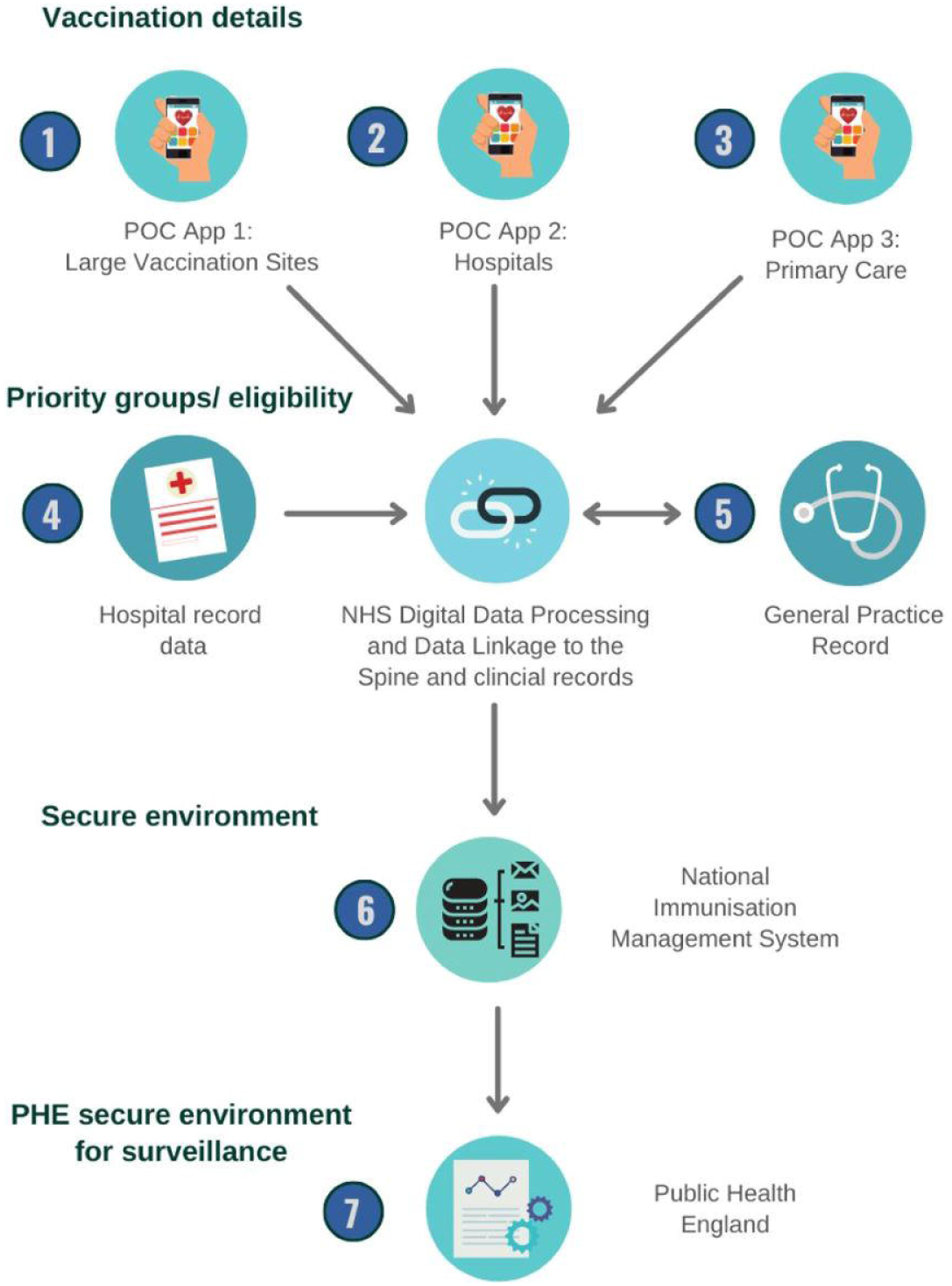

The data from NIMS is provided to PHE in two files;

a. A population denominator file of all persons that have an NHS number with accompanying demographic details, including ethnicity as defined in the 2001 census and 2011 census^9^. Individuals that are clinically extremely vulnerable are flagged based on information from their clinical record, and healthcare and social care workers are flagged based on NHS Electronic Staff Records.
b. A vaccination events file of all persons that have had a minimum of one COVID-19 vaccine recorded with accompanying details of the vaccination event, including location of where the vaccine was delivered, date the vaccine was administered, manufacturer information and vaccine batch number.

PHE import a daily feed of data into a SQL Server using automated SQL Server Integration Services (SSIS) jobs and stored procedures developed by PHE data engineers. Data cleaning and assurance is carried out in the staging area prior to the data being published to a data store. PHE deduplicates records, validates the NHS numbers using an NHS number check digit and assesses the data for any anomalies. Individuals must have a valid NHS number to be linked to the individual file with the persons demographics.

PHE set rules and derive a series of variables depending on the purpose the data is being used for. Vaccination details such as the dosing information and vaccination manufacturer are provided in Systematised Nomenclature of Medicine Clinical Terms (SNOMED CT)^10^. Vaccine batch numbers are either scanned (using a barcode scanner) or manually entered in the PoC apps. PHE undertake initial data cleaning of batch numbers, and then assign the vaccine manufacturer to individual records. Where the batch number provided is incomplete or blank, the SNOMED CT code is used to allocate the manufacturer.

For vaccine coverage purposes, individuals that received a second dose of the vaccine less than 20 days after their first dose are considered as not having a complete course of the COVID-19 vaccine, and only their first dose is used. For vaccine effectiveness and vaccine safety purposes all doses recorded in the NIMS are retained. Individuals recorded as both having been vaccinated and having refused/declined a vaccine were not recorded as having a vaccine as it is not possible to guarantee that the vaccine has been administered.

Records are then linked to individual postcodes from the GP record to allocate the region each individual resides in, the 2011 ONS rural/urban classification, and the 2019 Index of Multiple Deprivation (IMD) decile^11^. NHS numbers are used to link to additional datasets such as care home resident lists (maintained by NHS England and provided monthly), COVID-19 community level testing data, and COVID-19 related hospitalisations and deaths^12^.

### Data validation

As part of PHE’s role to monitor the COVID-19 pandemic, case-based enhanced surveillance system has been set in place to follow-up individuals who have tested for SARS-CoV-2. A questionnaire is administered by nurses over the phone to a random selection of individuals in England and asks for their vaccination details. We compared the self-reported vaccination dose, manufacturer and vaccination date from the enhanced surveillance system to the NIMS data on 24 August 2021.

## Results

The development of NIMS as a single national immunisation register has reduced the risk of inaccurate immunisation records for the entire population and the PoC apps provide a more consistent template of entering data across the country. The barcode scanners have reduced data entry errors and increased the completeness of individual level information for national vaccination programmes.

The NIMS has been highly beneficial for public health agencies, NHS England and Improvement and PHE, in supplying, delivering and evaluating the COVID-19 vaccination programme. Furthermore, this centralised system records details on vaccinations from across the country, delivered in all types of settings in quasi real-time. PHE receives daily outputs from NIMS allowing for rapid monitoring of trends in age-specific vaccine coverage and identifying sub-populations with poor coverage to be targeted quickly. Furthermore, PHE have been able to link COVID-19 testing data to NIMS for rapid vaccine effectiveness surveillance among defined cohorts, which historically has not been easy to conduct^13 14^. In the past, vaccine effectiveness studies for infections such as influenza have relied on a variety of data sources such as sentinel surveillance systems data, swabbing forms or data recorded in GP datasets obtained by telephone or postal letters^15 16^. Vaccine effectiveness for childhood immunisations have used aggregated national coverage estimates and local CHIS data derived from multiple sources and requires several user permissions and the linkage of several datasets^17-19^.

Finally, the NIMS data is retained in a highly secure server environment shared directly within PHE under The Health Service (Control of Patient Information) Regulations 2002; specifically regulation 3(1)(d)(iii) and (iv) – “monitoring and managing the delivery, efficacy and safety of immunisation programmes and adverse reactions to vaccines and medicines” ^20^, and under Section 251 of the 2006 National Health Service Act. This allows the common law duty of confidentiality to be set aside in specific circumstances where anonymised information is not sufficient, such as monitoring adverse reactions to vaccines and the delivery of efficacy and safety of immunisation programmes 21. PHE have a Data Sharing Agreement (DSA) which aligns with current Caldicott Principles 22 and sets a legal framework for using the data within the organisation. Those who wish to use the data must meet the DSA principles and apply for access to the data with an acceptable explanation of how the data will be used. The data are then shared in a secure SQL environment and all work is conducted on a secure server.

### Data validation

A total of 1,129 individuals had responded to the enhanced surveillance questionnaire by 24 August 2021. Of these individuals, a total of 309 of 316 (97.8%) of individuals were recorded in the NIMS as unvaccinated compared to those who self-reported as unvaccinated. A total of 813 of 813 (100.0%) of individuals were recorded in the NIMS as having at least one dose of the COVID-19 vaccine compared to those who self-reported as having one dose, of which 297 of 299 (99.3%) of individuals were recorded in the NIMS as having two doses of COVID-19 vaccine compared to those who self-reported having two doses.

Of the 813 people who were recorded in the NIMS as having at least one dose of the COVID-19 vaccine, 803/813 (98.8%) had the same manufacturer information for their first dose as that which was self-reported in the enhanced surveillance questionnaire while 297/299 (99.3%) people who were recorded in the NIMS as having had two doses of the COVID-19 vaccine self-reported the same manufacturer information.

Of the 813 people who had a first vaccination according to both the NIMS and the enhanced surveillance questionnaire, 799/813 (98.3%) self-reported a vaccination date for their first dose. There was good agreement between the vaccination dates; 675/799 (84.5%) had the same date of vaccination according to the NIMS and the enhanced surveillance questionnaire while 729/799 (91.2%) people self-reported a date which was within 3 days of the date recorded in the NIMS (median difference: 0 days, IQR: 0-0 days). Of the 299 people who had a second vaccination date recorded in the NIMS, 291/299 (97.3%) self-reported a vaccination date for their second dose. A total of 259/291 (89.0%) self-reported the same date as that which was recorded in the NIMS whilst 271/291 (93.1%) self-reported a date which was within 3 days of the date recorded in the NIMS (median difference: 0 days, IQR: 0-0 days) (Figure 2).

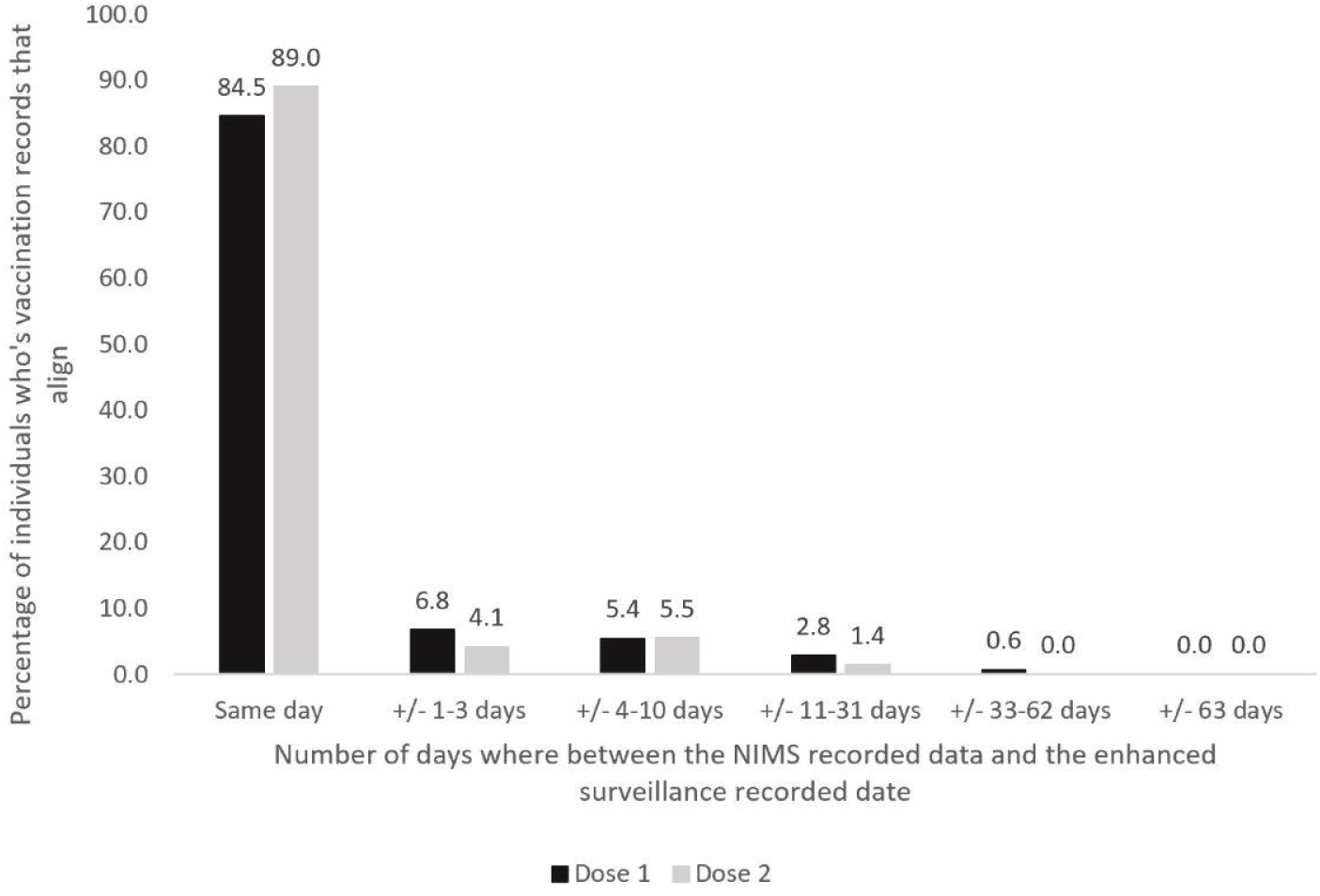

## Discussion

### Limitations and improvements to NIMS

Currently the data in NIMS can only be linked for individuals with a NHS number where demographic details can be populated from the GP record or hospital record data. Those without an NHS number can opportunistically receive a vaccine and a NHS number will be allocated upon vaccination. These individuals’ records will only contain information from their vaccine event. As such, it is possible that some people are missing opportunities to be invited for a vaccination. Though the numbers of individuals without an NHS number is marginal, the most vulnerable populations (migrants, asylum seekers) are more likely to not have an NHS number, and as such excluding them from the benefit of the NIMS may exacerbate health inequalities.

The use of PoC apps has made for more efficient data entry, however, errors due to manual entry are still possible. Many of these errors can only be validated at the vaccination site where the vaccine was delivered. The packaging of current vaccines do not leave sufficient space for barcodes to be added to individual vials and on some occasions barcodes have been pre-loaded at vaccination sessions in attempt to eliminate data entry error.

Finally, though the NIMS is based on a unique NHS number, this number is only linked to health services datasets. Linking to non-health related indicators such as occupation, emigration are not possible using the NHS number. Parts of the NIMS’ data have attempted to overcome this limitation by linking to other datasets such as the NHS Electronic Staff Record to identify substantive staff employed by the NHS which is only a subset of frontline healthcare workers. Some countries, such as Denmark, have overcome this barrier as they can use the a Civil Registration System (CPR-registeret) which also collects and updates information on non-health related factors that could affect vaccination uptake such as civil status, emigration etc^23 24^.

### Conclusions

Prior to NIMS, England did not have a national immunisation information system with individual level data for the entire population. Routine immunisations are recorded on GP records and in the CHIS for childhood immunisations up to the child’s 18^th^ birthday and work is being conducted on improving child health data through the Digital Child Health Transformation Programme^25^. Though these systems are capable of monitoring vaccination and sending invitations and reminders for vaccination appointments like the NIMS, as CHISs and GP systems are currently not all interoperable and risk duplicating or missing individuals, thus impacting accuracy of the data^3^.

The 2009 Swine Flu pandemic highlighted the importance of vaccine registers and immunisation information systems ^26^. The NIMS has served as a foundation for COVID-19 vaccines surveillance since the introduction of the COVID-19 vaccination programme and had begun being used for seasonal influenza vaccines. Without such a system centrally collecting individual level data we would not be able to provide vaccine coverage data with the same granularity and accuracy or provide some of the world’s first vaccine effectiveness estimates.

Furthermore, the results from validating NIMS data to self-reported vaccination details from the enhanced surveillance system indicates high accuracy and completeness of the data in the NIMS. It is important to note that the self-reported questionnaire is not a gold standard therefore the difference in complexness and accuracy observed may also be due to a lack of accuracy in the questionnaire results, rather than the NIMS. For example, recall bias or data entry errors in the self-reported forms could also account for some differences between the two systems. Any data quality issues in the NIMS can be flagged with NHS Digital who review the queries and amend the data when possible.

To appropriately monitor the COVID-19 pandemic globally and the impact of COVID-19 vaccines, countries around the world should adopt national immunisation registers of systems that can be rapidly linked to health data. In England, the NIMS should continue to develop to enable individuals with no NHS numbers to be monitored and expand to allow for linkage to non-healthcare related datasets that may impact access and behaviours to vaccines. Finally, the expansion of the NIMS to monitor other routine immunisation programmes would be highly valuable.

## Data Availability

Data may be available upon request.

## Conflict of interest

Funding from Public Health England for the submitted work; no financial relationships with any organisations that might have an interest in the submitted work in the previous three years, no other relationships or activities that could appear to have influenced the submitted work.

## Ethics Approval

Surveillance of covid-19 vaccination data is undertaken under Regulation 3 of The Health Service (Control of Patient Information) Regulations 2002 to collect confidential patient information (www.legislation.gov.uk/uksi/2002/1438/regulation/3/made) under Sections 3(i) (a) to (c), 3(i)(d) (i) and (ii) and 3(3).

## Funding statement

There was no external funding for this study.

## Acknowledgements

We would like to thank colleagues from System C & Graphnet Care Alliance, NHS Digital and South Central and West CSU for their continued support in providing the NIMS data that we needed for surveillance and monitoring vaccine coverage. We would also like to thank the COVID Data Store and ImmForm teams at Public Health England who process the daily NIMS feed at PHE. Finally we thank the PHE nurses.

## Notes

### Competing Interest Statement

The authors have declared no competing interest.

### Author Declarations

Surveillance of covid-19 vaccination data is undertaken under Regulation 3 of The Health Service (Control of Patient Information) Regulations 2002 to collect confidential patient information (www.legislation.gov.uk/uksi/2002/1438/regulation/3/made) under Sections 3(i) (a) to (c), 3(i)(d) (i) and (ii) and 3(3). Ethics approval also been approved by the head of research governance at Public Health England

